# Drug company payments to Australian healthcare professionals

**DOI:** 10.1101/2025.09.10.25335461

**Authors:** Malcolm Forbes, Kane Harvey, Spencer Schien, Ashleigh Hooimeyer, Adrian Pokorny

**Affiliations:** Mental Health, Drug and Alcohol Services, Barwon Health and the Institute for Mental and Physical Health and Clinical Translation (IMPACT), School of Medicine, Deakin University, Geelong, Victoria, Australia; Mental Health & Wellbeing Service, Mercy Health, Melbourne, Australia; Rayshade Consulting, Milwaukee, Wisconsin, USA; School of Pharmacy and Charles Perkins Centre, Faculty of Medicine and Health, University of Sydney, Sydney, Australia; Chris O’Brien Lifehouse, Sydney, Australia

**Keywords:** Pharmaceutical industry, Biopharmaceutical industry, Drug company, Financial payments, Conflicts of interest, Competing interests, Medical ethics

## Abstract

**Objectives:** To map specialty-specific industry payments to Australian healthcare professionals over a nine year period (2015–24), quantify distribution and concentration of payments, and compare patterns with those reported internationally.

**Design:** Repeated cross-sectional and cohort analysis

**Setting:** All public disclosures of payments by pharmaceutical companies to Australian healthcare professionals, 1 October 2015 to 31 October 2024.

**Participants:** 23,528 named healthcare professionals who received at least one payment from a Medicines Australia member company.

**Main outcome measures:** Primary outcomes were: (1) payment volume (number and value of payments, distribution by purpose of payment); (2) profession and specialty reach (proportion of registered practitioners with ≥ 1 payment; and (3) concentration of payments (the share of the total value received by top 1%, 5%, and 10% of recipients). Secondary outcomes were persistence of payments across reporting periods, breadth of company relationships, and company-specialty payment patterns.

**Results:** 104,663 payments with a total value of A$164,445,101 (£79.0m) were reported. Payments ranged from A$1 to A$114,400, with a median of A$1,000 (IQR A$582 to A$1,600). Over 80% of paid clinicians received at least one payment for attending an educational meeting and over 90% of the total value of funding covered travel or fees for service. Medical practitioners accounted for A$148.7m (90.4% of total spending); nurses received A$11.7m (7.1%), and pharmacists A$1.8m (1.1%). In total, 12.2% of doctors, 2.1% of pharmacists, and 1% of nurses in Australia received at least one payment between 2015 and 2024. Specialist reach was highest in clinical haematology and oncology (86%) and rheumatology (82%). Payments were highly concentrated: the top 1% of clinicians received 25% of all dollars, the top 5% 55.3%, and the top 10% 70.5%. Persistence of payments was common, with 30.3% of clinicians appearing in more than two reporting periods. Annual totals peaked in 2016/17 (A$30.8m), contracted between 2018/19 to 2021/22 and rebounded by 2023/24 (A$21.6m).

**Conclusion:** Pharmaceutical company payments to Australian healthcare professionals were common and highly concentrated, particularly in specialties with current on-patent medicines, mirroring patterns reported in the United States. Competing interests management should prioritise independence in clinician education, avoid company sponsored drug promotion, and maintain robust disclosure and governance.

**What is already known:** Industry payments to clinicians are common internationally and can influence prescribing. Australian evidence to date has been fragmented by period, profession, or specialty.

**What this study adds:** Using all named disclosures from 2015–24, payments in Australia were widespread, with over 1 in 10 doctors in Australia receiving at least one payment. Payments were also highly concentrated, with the top 1% of recipients receiving 25% of the total payment amount. Haematology and oncology and rheumatology had the highest specialty reach. Findings support stronger competing interests management in medical education and continuing professional development.

## INTRODUCTION

In his 1973 report on the aggressive marketing of amphetamines and barbiturates in the USA, American journalist John Pekkanen observed that “the doctor is feted and courted by drug companies with the ardour of a spring love affair. The industry covets his soul and his prescription pad because he is in a unique economic position; he tells the consumer what to buy” (1). In the ensuing half century, pharmaceutical company promotion and marketing has matured into a trillion-dollar enterprise (2). Payments are made for consulting, speaking at conferences, advisory board participation, travel to educational meetings, conference registration, accommodation, food and drink, and research activities (3). A systematic review conducted in 2021, encompassing 36 studies, identified a positive association between pharmaceutical payments and prescribing rates. The analysis found a temporal relationship and dose-response effect, supporting the inference of a potential causal link (4). Increasing entanglement between industry and academic medicine led a former editor in chief of the New England Journal of Medicine to conclude that, “it is simply no longer possible to believe much of the clinical research that is published, or to rely on the judgement of trusted physicians or authoritative medical guidelines” (5).

Transparency schemes have emerged to address these concerns, but their scope varies by jurisdiction. In the United States, the Physician Payments Sunshine Act created the Open Payments database in 2013, which records payments beyond a nominal value to all healthcare professionals (6). The United Kingdom, in contrast, has no statutory disclosure law and instead relies on an industry-led voluntary scheme. The Disclosure UK database, until recently, allowed individual clinicians the opportunity to remain anonymous. In Australia, Medicines Australia, the industry body representing most pharmaceutical companies, has mandated reporting of all payments to healthcare professionals since 2015, except for food, beverages, and research. A centralised database was created in 2019 (7).

To date, published analyses of pharmaceutical payments to healthcare professionals in Australia have been limited and only one study has examined the distribution of payments to all medical specialties (7). Previous investigations have examined aggregate spending or focused on limited time periods. Few studies have reported payments to non-medical clinicians in Australia (8). To fill this gap in the literature, here we provide the most comprehensive analysis of payments to Australian healthcare professionals conducted to date.

## METHODS

### Study design and data sources

We conducted a repeated cross-sectional and cohort analysis of all transfers of value from pharmaceutical companies to Australian healthcare professionals, from 1 October 2015 to 31 October 2024. The period spans the introduction of enhanced transparency requirements. Six-monthly reports were obtained from the University of Sydney repository (9) and, from 2019, the Disclosure Australia portal (10). Each record included company, recipient name, professional type, practice address, descriptors of service and event, the category of payment recipient (individual, employer, or third party), itemised amounts (registration fees, travel, and fees for service), and the time period during which payment occurred. Disclosure Australia information does not list the specialty of the payment recipient, so further data linkage was required. Payments to organisations and research-related transfers were outside the scope of this study. Further details are found in the Supplementary Methods.

### Outcomes

Primary outcomes were: (1) payment volume, defined as the total value in nominal Australian dollars (A$) and number of payments over the reporting period, classified by service type; (2) profession and specialty reach, defined as the count and proportion of practitioners from each professional group (and for medical practitioners, specialty field) who received at least one payment over the study period, using Australian Health Practitioner Regulation Agency (AHPRA) registration data to link healthcare professionals listed by Medicines Australia to their specialty; and (3) payment concentration, quantified as the share of total dollars received by recipients ranked by cumulative payments.

Secondary outcomes included persistence of payments, defined as the proportion of clinicians appearing in multiple reporting periods; breadth of company relationships, measured as the number of distinct companies making payments to each clinician; and descriptive analyses of time trends by calendar year. We also computed company-level distributions, ranked by total spend, and built company-specialty matrices.

### Statistical analysis

For each specialty we reported the total value of payments, the number of practitioners who received at least one payment, and specialty reach using 2023/24 AHPRA registration data (11). Reach was defined as the percentage of registrants with ≥ 1 payment. Per person payments were described using the median and interquartile range. Payment concentration was quantified as the share of total dollars received by the top 1%, 5%, and 10% of recipients, ranked by cumulative payments, with winsorisation used to exclude outliers as a sensitivity check. A Gini coefficient was calculated to provide a summary measure of overall payment inequality.

For specialty reach, given not all companies are represented in our database, 95% confidence intervals were calculated using Wilson binomial intervals. These intervals do not capture potential under-reporting. Company–specialty cross-tabulations were created for the entire period, and for each specialty, the share of payments received by the leading company was reported. Time trends were analysed across 12-month periods. All monetary values were expressed in nominal Australian dollars, except in Figure 4, which is inflation-adjusted to 2023/24 AUD. Analyses were conducted in R (version 4.5.0). Further details are provided in the Supplementary Methods.

### Ethics

The University of Sydney Human Research Ethics Committee determined the study to be exempt from review because it used publicly available data.

### Patient and public involvement

While we appreciate the importance of patient and public involvement in research, we did not have the requisite funding or resources to support this activity. Thus patients and members of the public were not involved in the design, conduct, reporting, or dissemination of this research.

## RESULTS

### Payment volume

Between 1 October 2015 and 31 October 2024, Medicines Australia member companies reported 104,663 payments to 23,528 individuals from 45 companies, totalling A$164,445,101. Payments ranged from A$1 to A$114,400, with a median payment of A$1,000 (IQR A$582 to A$1,600) (Supplementary Figure S1). Four in five clinicians received at least one payment for attending an educational meeting (Table 1). The majority of payments funded travel or fees for service.

**Table 1:** Payments by service provided.

| Service | Total payments (A\$)<br>(n = 104,663) | Proportion of all clinicians<br>who received ≥ 1 payment<br>(%)*<br>(n = 23,528) |
| --- | --- | --- |
| Educational meeting attendee | 70,545,405 | 81 |
| Educational meeting speaker or<br>chairperson | 52,657,348 | 32 |
| Advisory board committee<br>attendee | 24,946,725 | 15 |
| Consultant | 11,664,027 | 11 |
| Other | 4,631,596 | 7 |

*\*A clinician can receive multiple payments for different services, so percentages do not sum* *to 100%.*

### Profession and specialty reach

Medical practitioners received A$148.7 million (17,519 recipients; 90.4% of total spending), nurses A$11.7 million (4,611; 7.1%), and pharmacists A$1.8 million (807; 1.1%). Allied health and Other categories together accounted for A$2.2 million (1.4%). Using AHPRA 2023/24 registrant counts, 12.2% of all medical practitioners, 2.1% of all pharmacists, and 1% of all nurses received at least one payment within the study window.

Among medical practitioners, industry payments were concentrated in specialties where prescribing involves on-patent or competition-limited medicines. Aggregate spending was highest in cancer and cardiovascular care (Figure 1). General practice ranked third by total value, reflecting the large number of doctors in this specialty rather than high per-clinician payments (Figure 2). Surgical and critical care disciplines attracted relatively few payments.

**Figure 1.**
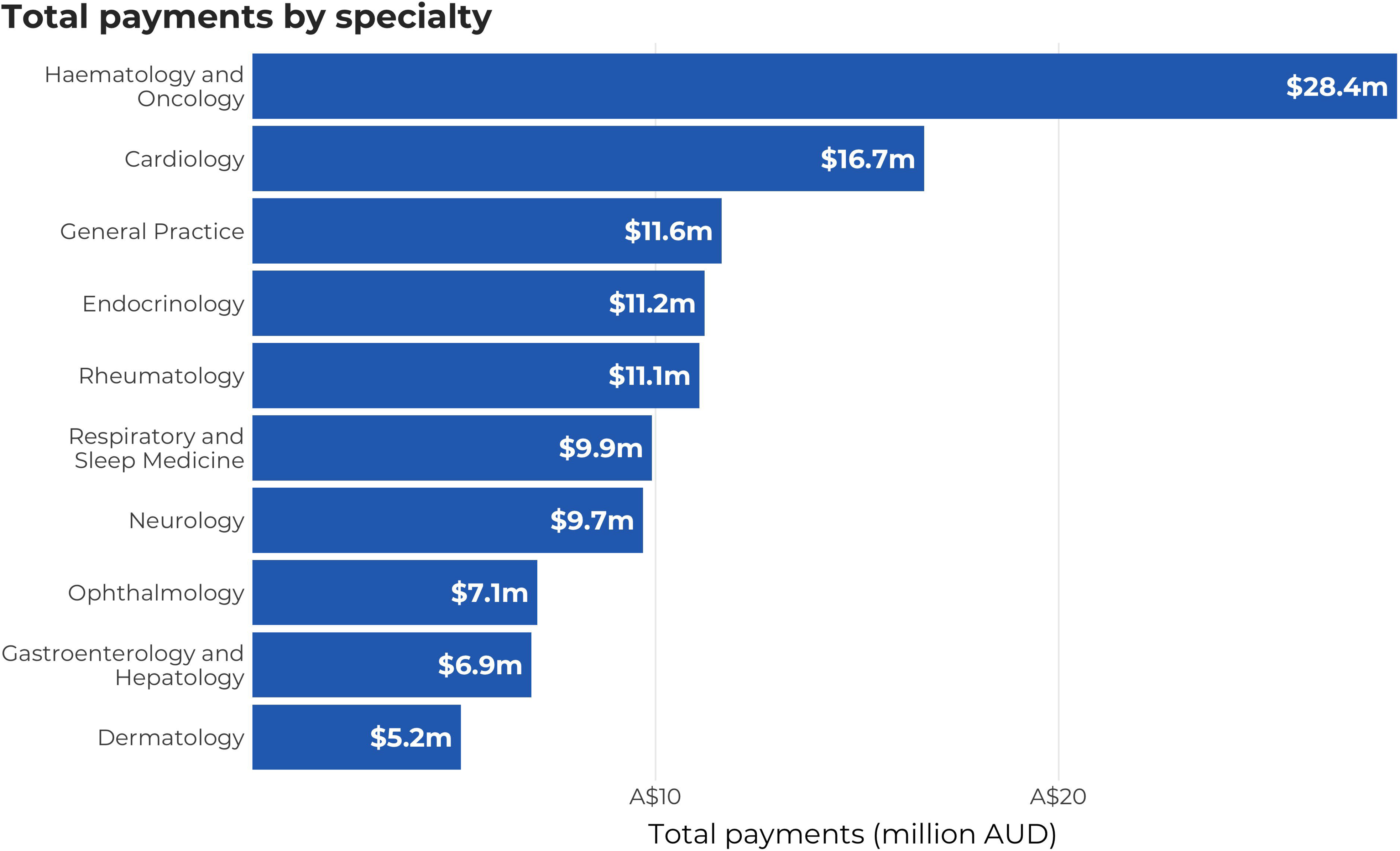

**Figure 2.**
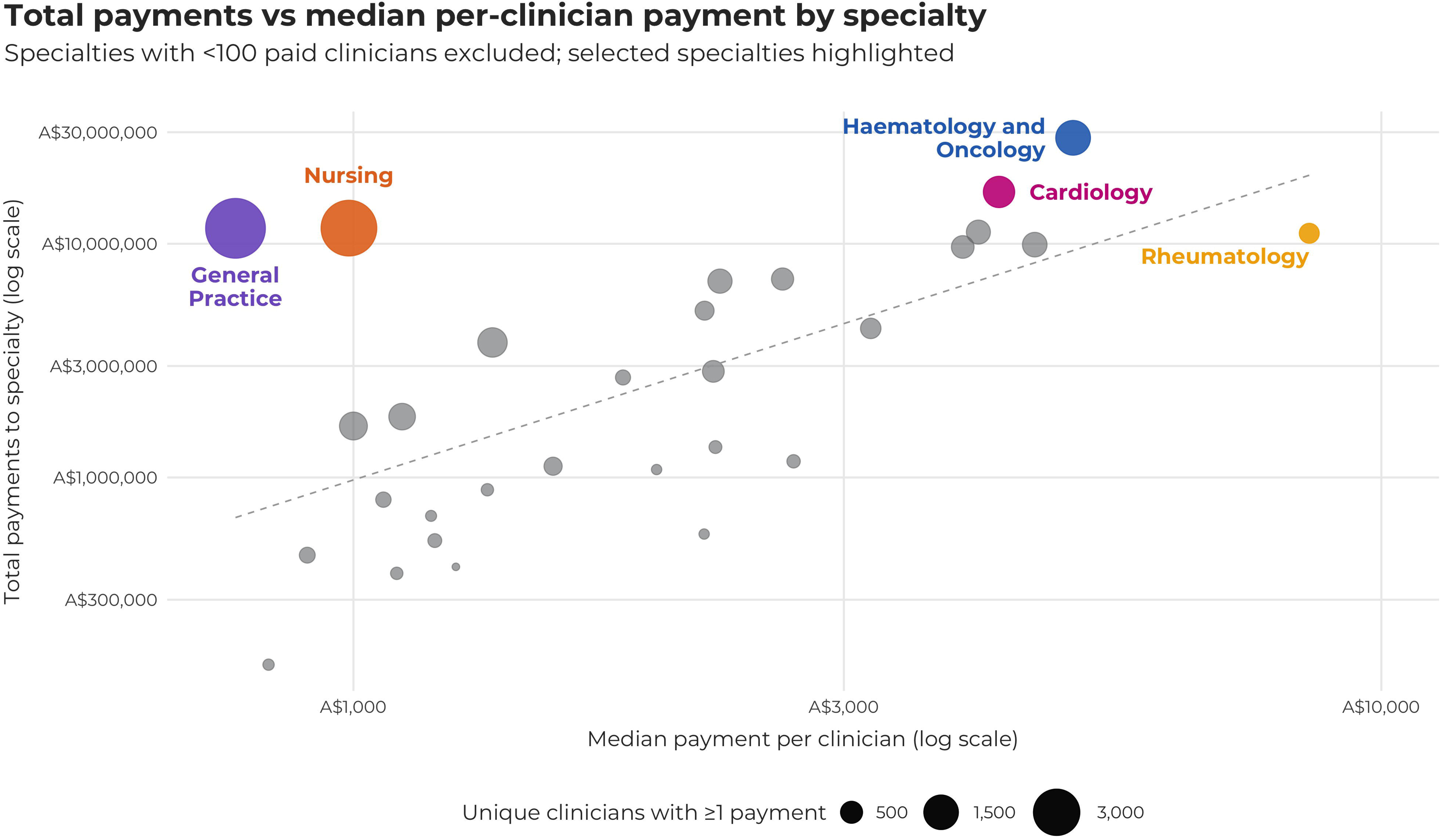

Specialist reach was highest in clinical haematology and oncology (86%) and rheumatology (82%) (Figure 3). This figure was restricted to specialties with more than 150 fellows (see Supplementary Methods).

**Figure 3.**
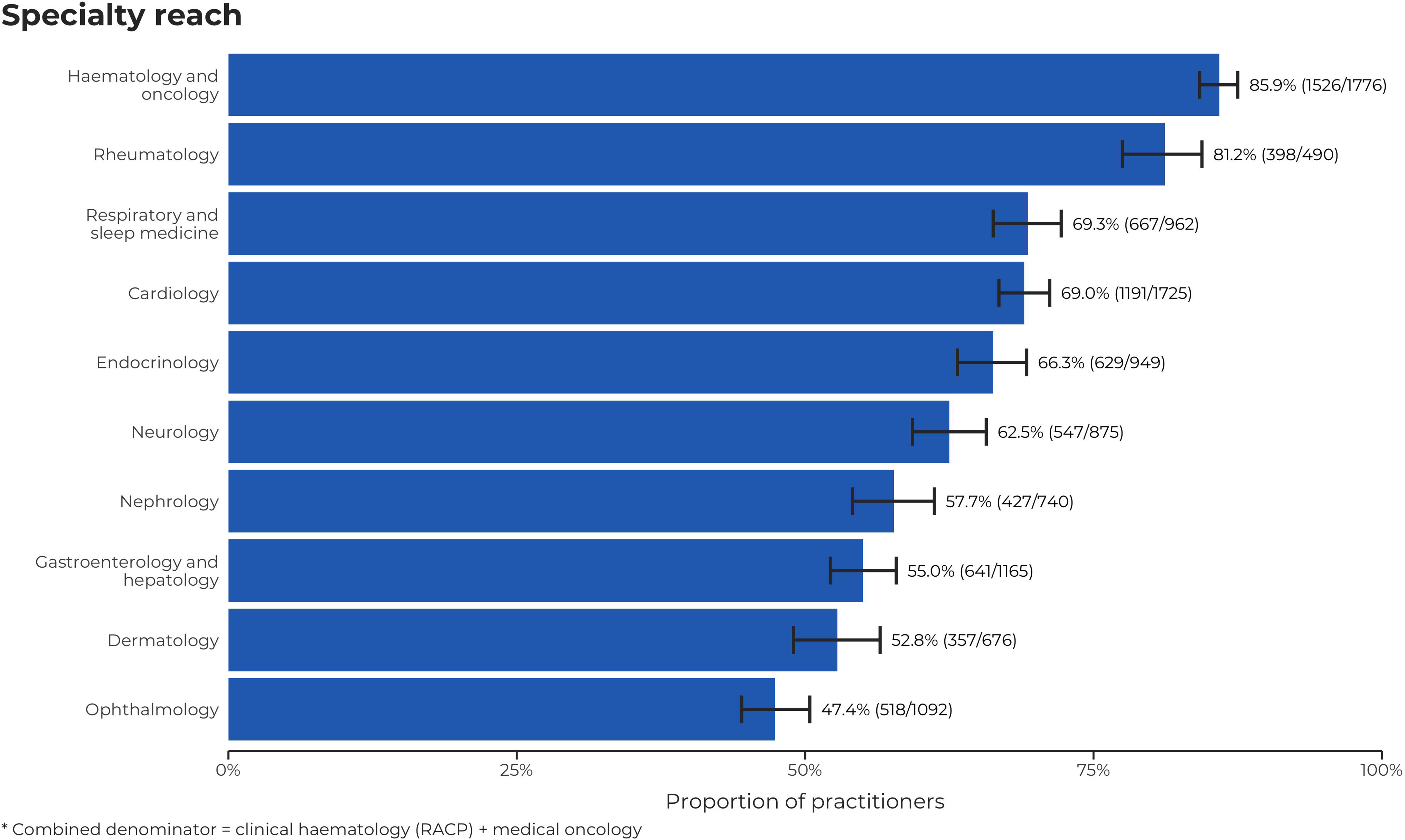

### Payment concentration

Total payment amounts were concentrated among a small number of recipients, with the top 1% (n = 236) accounting for 25% of total dollars; the top 5% (n = 1,180) for 55.3%; and the top 10% (n = 2,359) for 70.5% (Supplementary Figure S2). Winsorisation confirmed the robustness of this concentration. When payments above the 99th percentile were capped, the top 1% accounted for 16.3% of all dollars, while the top 5% and 10% still accounted for 50.0% and 67.1%, respectively. With more stringent caps at the 95th and 90th percentiles, the top 10% of clinicians continued to receive over half (56.0%) and nearly half (42.7%) of total payments, respectively. Review of the public profiles of individuals in the top 1% by dollar value revealed they were senior specialists, usually with university and specialty college appointments. Among paid clinicians, 30.3% had payments recorded in more than two reporting periods, 20.8% in more than three, and 15.4% in more than four. Persistence across greater than 2 periods was most common in general practice (n = 957), clinical haematology and oncology (n = 954), nursing (n = 942), cardiology (n = 640), and respiratory medicine (n = 406). The median number of distinct companies per clinician was 1 (IQR 1–2). The top 1% by company count (≥ 9 companies) were disproportionately drawn from clinical haematology and oncology, rheumatology, cardiology, endocrinology and gastroenterology.

### Payments across the study period

Annualised totals by reporting year showed a peak in 2016/17, a contraction in 2017/18, a marked reduction in 2018/19, and a subsequent rebound in 2022–2024. The number of unique recipients per year followed a similar pattern, consistent with reduced activity during the COVID-19 period and later recovery. These trends are reflected in the trajectories of the five largest paying companies (Figure 4). While COVID-19 contributed to a reduction in payments from 2020-2023, the 2018/19 drop likely reflects the transition to a centralised database, with incomplete coverage of payments within the database, subsequently addressed by Medicines Australia guidance (12). Some companies also ceased membership of Medicines Australia over the study period and thus were not compelled to disclose payments thereafter.

**Figure 4.**
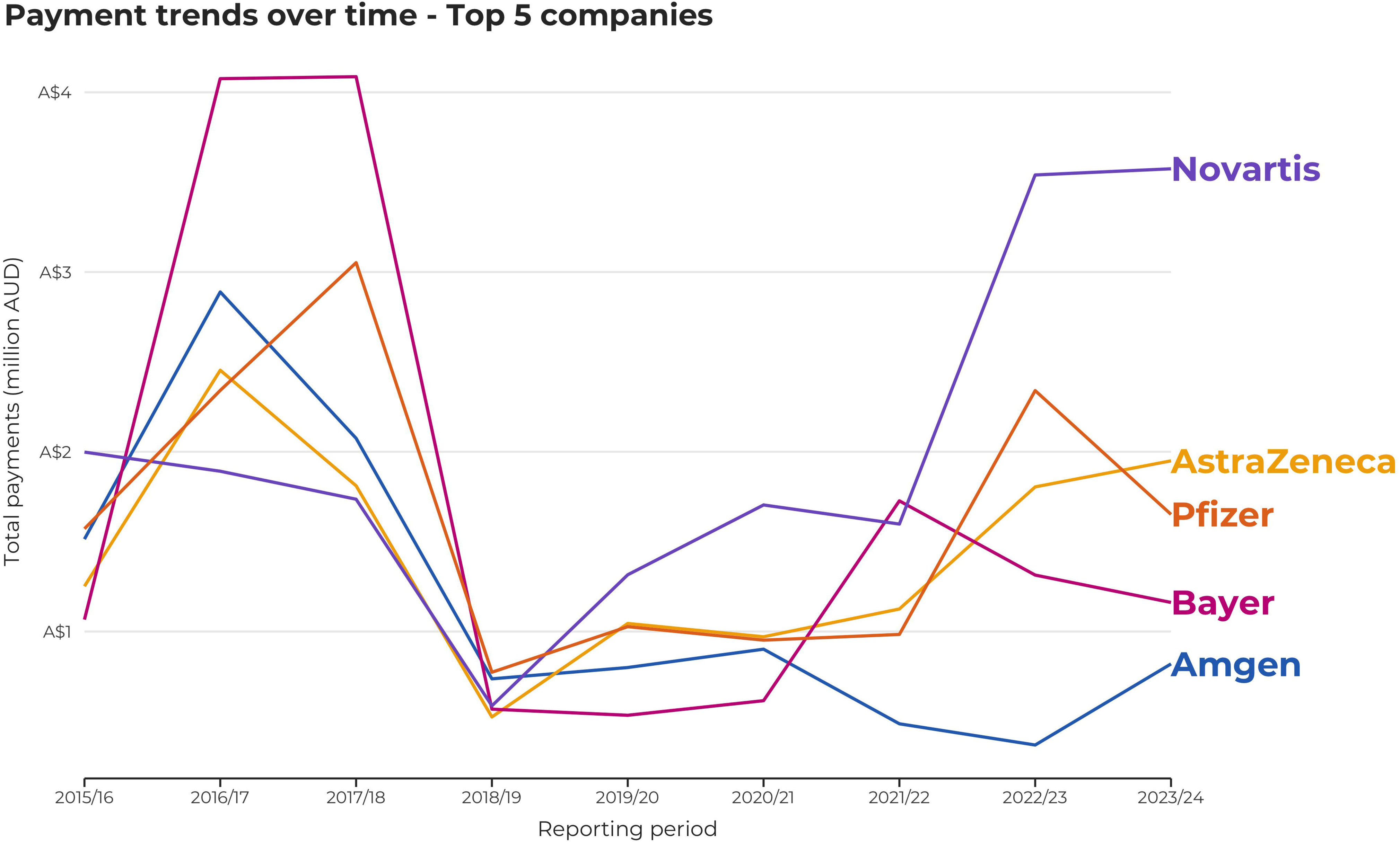

### Further analyses

Data checks revealed that 1,186 individuals had at least one duplicate entry, amounting to A$ 3,309,739. Excluding exact duplicate rows, the total payment between 2015 and 2024 would be A$ 161,135,362. Review of original company reports available over the study period noted duplicates were not data errors and were thus retained in the dataset. Duplicate payments may occur for a range of reasons, including if a company has hired a clinician to give talks and paid the same honorarium per speaking engagement. Further analysis, including company-level distributions and company-specialty relationships are detailed in the Supplementary Results.

## DISCUSSION

Our findings show that across nine years of industry mandated disclosures, there were 104,663 pharmaceutical company payments to 23,528 healthcare professionals totalling A$164.4m, with the top 1% of recipients accounting for 25% of the dollar value of all payments. Payments were strongly clustered in specialties managing on-patent medications, with haematology and oncology (86%) and rheumatology (82%) showing the highest reach.

### Possible explanations and implications

Pharmaceutical companies in Australia appear to follow a two-track strategy in their payments to healthcare professionals: broad but shallow engagement, with many specialists receiving one-off payments; and a deep, focused investment in a cadre of influential clinicians, often termed key opinion leaders. Key opinion leaders (KOLs) are highly connected physicians who often hold leadership positions within medical associations (13, 14) and operate as influencers within their professional communities (15). US evidence quantifies why companies prioritise payments to KOLs: after payment, prescribing rises among both paid physicians and their peers. Over a three year study period, about one-quarter of the total increase in novel anticoagulant prescription volumes came from peer spillover effects (16). These dynamics make sense of our finding of a heavy concentration of payments among a minority of medical practitioners. This concentration also occurred among nurses and pharmacists, albeit to a lesser extent. While nurses and pharmacists typically do not prescribe in Australia, they can serve as conduits to prescribers and may influence patient attitudes toward medications (8).

Payments were concentrated in cancer and other high-cost therapeutic areas. In clinical haematology and oncology, most specialists had received at least one payment over the study period. This may reflect perceived benefits of industry relationships, including access programmes and pragmatic support for rapidly evolving treatments (17). Among cancer physicians, there also appears to be recognition that some industry engagement may improve patient outcomes, such as advisory board membership to help design suitable clinical trials (17).

### International comparisons

Our findings build on a systematic review that found up to 63% of cancer physicians in the United States receive payment from or maintain financial ties with industry (18). Similarly, in Japan 71% of consultant rheumatologists received a payment between 2016-2019 (19). We showed that over a nine year period, an even higher proportion of Australian cancer physicians and rheumatologists received payments.

International research has found that payments are significantly concentrated among KOLs, including clinical practice guideline authors and medical journal editors (18, 19). Our concentration measures mirror patterns seen in analyses of US Open Payments, though absolute totals are smaller in Australia because the industry self-regulatory scheme reports a narrower set of categories (fees for service, travel, and registration), excludes research payments, and does not require reporting by non-member companies. By contrast, US Open Payments includes general payments, research payments, and payments from both pharmaceutical and medical device companies (20).

International comparisons suggest that, despite improvements since 2015, Australia’s transparency regime still falls short of best practice (6). The current scheme is self-regulatory, covers a narrow set of payment types, and does not systematically report research payments to individual clinicians. It misses food and beverage payments, despite evidence suggesting these payments influence prescribing behaviour (21) and are present in more than 90% of industry sponsored events in Australia (22). Its financial penalties for under-reporting are comparatively small (23). A comprehensive, legislated model for transparency reporting could require all pharmaceutical and medical device companies to report all significant transfers of value using unique clinician identifiers (e.g. AHPRA registration number). Legislation could also require records remain public for longer than 3 years, and publish a single, searchable public database that uses fuzzy name matching. Such a registry would enable independent scrutiny and support stronger competing interests policies by professional bodies and journals.

### Strengths and limitations

This study provides nine years of national, individual-level data across multiple healthcare professions. It is the most comprehensive Australian analysis conducted. Previous Australian research has focused on doctors alone or on event-level disclosures (7, 24). The study is strengthened by quantification of payment inequality and use of winsorisation to assess payment concentration. However, there are several important limitations. First, the work relies on a self-regulatory transparency system which is vulnerable to gaps in adherence and reporting requirements. A recent analysis of The Association of the British Pharmaceutical Industry database found deficiencies in the information provided (25) and previous research has revealed data quality issues in Medicines Australia payment reports (26). Second, the payment disclosures omit non-Medicines Australia member companies so under-ascertainment is likely and may vary by specialty. Third, some misclassification of specialty is likely as Medicines Australia does not provide an individual’s unique clinician identifier (26) and AHPRA does not require healthcare professionals to practice under the registered name (27). Despite manual checks and a low misclassification rate, greater residual error in specialty classification remains a possibility. Furthermore, because searches were conducted up to 10 years after some payments were made, there is potential for misclassification if healthcare professionals completed specialty training or changed their profession or specialty between the payment date and the time of search. Finally, except in Figure 4, we did not adjust for inflation over the study period, meaning that earlier payments therefore have higher purchasing power than later payments.

### Professional practice and policy implications

In debates over the appropriateness of receiving pharmaceutical company payments, there is a tendency to generate arguments that fit prior beliefs, a form of motivated reasoning (28): proponents of stricter limits may over-ascribe harm whereas clinicians comfortable with industry funding of healthcare professionals may over-ascribe benefit. Discussions around the probity of industry engagement often contain moral overtones of corruption versus purity, shaped by one’s perception of the integrity of pharmaceutical companies. A balanced view may be achieved by recognising there is robust evidence that documents industry harms and benefits. Harms include, inter alia, “disease mongering” that widens illness definitions to drive drug sales (29), the opioid crisis that was fuelled by unlawful drug promotion (30), and repeated payment of kickbacks to doctors (31). Benefits include the rapid development and rollout of an effective COVID-19 vaccine (32), immune checkpoint inhibitors (33), and novel HIV treatments (34), among others. As noted in a recent debate article, “the profit driven pharmaceutical industry is the worst system for discovering new drugs, apart from all the others” (35).

Some industry payments benefit patients (36); for example, companies may compensate clinicians for advisory board work on early-stage drug development at rates commensurate with ordinary professional income. However, these advisory board payments, which are sometimes used to assist submissions for public funding of medicines, constitute a minority of payments to Australian healthcare professions; most payments sponsor clinicians’ attendance at educational meetings. Over the past nine years, at least A$70.5 million was paid for this purpose, raising concerns about the independence of clinical education. By a simple syllogism, first outlined over 60 years ago – if (1) clinical decisions should be guided primarily by patients’ interests, and (2) the education shaping those decisions should be independent of parties with a financial stake – then it follows that treating physicians should not attend pharmaceutical company sponsored education (37).

Empirical findings are consistent with this conclusion. Industry sponsored education is associated with patient harms, including higher costs from lower rates of generic prescribing (38) and selective acknowledgment of adverse effects of promoted medicines (39, 40). In oncology, manufacturer payments correlate with the delivery of non-recommended or low-value care for some conditions (41). A recent analysis of US Medicare data found that marketing payments to cancer-treating clinicians were followed by increased prescribing and higher spending on the promoted drugs, with no detectable improvement in 12-month mortality after adjustment for cancer type, comorbidities, recent hospitalisations, and demographics (42).

Finally, our findings have implications for public trust in healthcare professions in Australia. Experimental evidence suggests physicians who receive high payments are rated lower on honesty and fidelity by patients (43). Supporting this finding, public disclosure of payments is associated with lower trust in one’s own physician, regardless of whether respondents knew their physician had received payments (44). This study found that in a subgroup of participants, a patient’s trust in their physician increased if they knew they had not received any industry payments. However, given the small sample size, caution should be exercised in interpreting this finding (45).

There is a need to understand why so many clinicians accept industry funding to attend educational meetings. A U.S. historical perspective suggests the issue is structural: as professional bodies and regulators relaxed or relinquished core responsibilities for postgraduate drug education, pharmaceutical firms stepped into the breach and, over time, were even encouraged to assume a central role in physician education (46). As Marcia Angell, former editor in chief of the New England Journal of Medicine, observed, companies have perpetuated a “gigantic fiction” that they are “not in the business of selling drugs but also in the medical educational business” despite fiduciary duties that orientate them toward sales (47). Unlike many countries, Australia had a unique public commitment to the education of healthcare professionals on medicine use, via the National Prescribing Service. This organisation received core government funding to provide a range of educational services for 24 years but was defunded in 2022 (48). While specialty colleges in Australia are heavily involved in the education of trainees, this does not exist to the same degree for consultant specialists. Absent a renewed commitment by independent and public bodies to steward postgraduate drug education, the vacuum may be filled by pharmaceutical companies, deepening promotional influence over prescribing.

## CONCLUSION

This study found that pharmaceutical company payments to Australian clinicians were common and highly concentrated, most often for educational meeting attendance. Patient care should be guided by best evidence rather than promotional influence, and patients rightly expect unbiased advice. Our findings should prompt clinicians – particularly those invited to sponsored education – to consider whether accepting payments serves patients’ interests. Clinicians, colleges, and policymakers should prioritise independent continuing education and strengthen competing interests management in guideline development. In contrasting the priorities of healthcare professionals and the priorities of pharmaceutical companies, we encourage readers to reflect on the words of former editor in chief of the New England Journal of Medicine, Dr Marcia Angell: “Drug companies are not charities; they expect something in return for the money they spend, and they evidently get it, or they wouldn’t keep paying” (5).

## Data Availability

All data produced in the present study are available upon reasonable request to the authors.

## DATA SHARING STATEMENT

Data are available on a public repository at https://github.com/malforbes/PharmaPayments. This repository contains raw data and analysis and visualisation scripts.

## AUTHOR CONTRIBUTIONS

- **Malcolm Forbes**: Conceptualisation; Methodology; Investigation; Data curation; Formal analysis; Visualisation; Writing – original draft; Writing – review & editing; Supervision; Project administration.
- **Kane Harvey**: Data curation; Investigation; Writing – review & editing.
- **Spencer Schien**: Formal analysis; Validation; Visualisation; Writing – review & editing.
- **Ashleigh Hooimeyer**: Data curation; Investigation; Validation; Formal analysis; Writing – review & editing.
- **Adrian Pokorny**: Validation; Writing – review & editing.

Dr Forbes is the guarantor, accepts full responsibility for the work and the conduct of the study, had access to all data, and controlled the decision to publish.

