## Supplementary Methods for "Drug company payments to Australian healthcare professionals"

**Data curation, quality control and analysis approach**

Data on payments to health professionals from November 2019 to October 2024 were downloaded from a centralised repository (1). This information included individual payments to health professionals, with columns of company name, name, healthcare professional type, practice address, type of service, type of event, payment recipient, registration fees, travel costs, fees for services, and reporting period. These were merged with legacy reports from October 2015 to October 2019 data obtained from the University of Sydney repository (2). We cross-validated overlapping 2019 entries between legacy files and the centralised database. For all payment values, it was assumed that all payments were in Australian dollars.

Previous exposure to Medicines Australia data has revealed errors in classification, with a small number of healthcare professionals listed under different names or with the incorrect profession or specialty (3). Thus in lieu of a semi-automated approach using the *stringdist* package in R (4), as outlined in Forbes et al. (5), manual searches were conducted for all individuals. These searches were completed by K.H., M.F., and A.H. between June 2024 and April 2025. Each name was entered into the AHPRA Register of Practitioners (6) using the default drop-down option. In cases where an exact match was uncertain (for example, a common name shared by multiple practitioners), we conducted further verification. This included cross-checking the practitioner’s details via hospital or clinic websites, conference speaker biographies, and other public sources to confirm the individual’s identity and specialty. Disagreements were resolved by consensus. The list of fields of specialties was recorded as per the List of Specialties, Fields of Specialty Practice and Related Specialist Titles (7). The only specialties that were combined were clinical haematology and medical oncology. Doctors-in-training were listed separately.

Given the large volume of searches required, we only recorded specialty and did not record other details about individual practitioners, such as year of fellowship or sex. There were 134 individuals for whom a specialty could not be ascertained. It may be the case that these individuals were registered under one name on AHPRA but received payment under another. Furthermore, there were some medical professionals who had more than one medical specialty. To prevent double-counting, we assigned each individual a primary specialty using online searches to identify their main area of work. Transfers of value may have occurred in their alternate specialty instead. There was also a delay between payment occurring and the healthcare professional being searched on AHPRA and there is potential for misclassification if healthcare professionals received a payment as a doctor-in-training but then completed specialty training or changed their profession or specialty between the payment date and the time of search. However, few medical practitioners change specialty, and most payments are directed toward senior practitioners rather than doctors-in-training, so any such misclassification is unlikely to materially affect our findings.

For each specialty we estimated pharmaceutical company reach (the proportion of registered clinicians in each specialty who received ≥ 1 payment) by linking payments to AHPRA 2023/24 workforce counts. Consistent with prior work (5), we combined clinical haematology and medical oncology denominators when reporting ‘clinical haematology/oncology’ reach. The number of specialists in each specialty field at the end of the study period was utilised to calculate proportions. This provides the most conservative estimate of pharmaceutical company reach. However, it does not take into account individuals who have retired over the study period and thus may artificially exaggerate proportions. The supplementary tables R3 and R9 can be found on the AHPRA website (8).

Data validation was undertaken throughout the data collection process. An initial data accuracy check occurred in January 2025, with K.H. completing checks to confirm the assigned specialties of the 300 individuals who received the highest payment amounts. A second data accuracy check was undertaken between January and April 2025 by A.H., with a random sample of 10% of the dataset. This revealed a specialty misclassification rate of 1.3%. These misclassifications were corrected. A final data accuracy check was conducted in July 2025 by M.F. to screen a further 10% of randomly selected practitioner records using an automated process. For each name and address pair, a Python script queried Google via the serper.dev API, retrieved the title of the first organic result, and matched this text against a curated dictionary of 70 specialty descriptors. The concurrence between manual and automated checks supports the overall integrity of the final dataset.

To express the time series in Figure 4 in 2023/24 AUD, nominal six month totals were deflated using the Australian Consumer Price Index, anchoring each payment to the end month of its reporting period (9).

Data cleaning occurred between July and August 2025 by M.F. Company names were mapped to standard labels (for example, both “Bristol-Myers Squibb” and “Bristol Myers Squibb” were recoded as Bristol Myers Squibb). Analyses were conducted at the brand level. We did not combine brands after mergers or acquisitions (e.g. Celgene to Bristol Myers Squibb; Shire to Takeda; Alexion to AstraZeneca; Actelion to Janssen; Vifor Pharma to CSL), and we treated spin-offs as separate entities (e.g., Alcon from Novartis; Organon from MSD). There were some companies who ceased to be members of Medicines Australia and thus stopped providing data on payments during the study period. This may have led some specialties to have spuriously low payment totals. Free-text fields describing service type, event type and payment recipient were collapsed into concise dictionaries – ten service categories, eight event categories and three recipient categories. In total, 6.3% of all payments were made to the healthcare professional’s employer or a third party. These records were retained, as such transfers may ultimately benefit the named clinician (for example, sponsored registration or support routed via an institution).

Formal data analysis was undertaken by M.F using R version 4.5.0 and checked by S.S. Data visualisation was undertaken by S.S. The dataset and all analytic code, including analysis and visualisation scripts, are available on a public repository. The cleaned analysis dataset provided for replication excludes the address field to protect privacy as some individuals appear to have listed their personal address instead of their professional address.

**SUPPLEMENTARY RESULTS**

**Companies**

Overall, Novartis, Bayer, and Pfizer were the three largest spenders, together accounting for 25.4% of all disclosed dollars (Table S1). Company rankings varied over time, consistent with product cycles and specialty launches.

**Table S1: Companies by total payment, top 10**

| **Company** | **Total payment (A$)** | **Share of total payments (%)** |
| --- | --- | --- |
| Novartis | 15,994,401 | 9.7 |
| Bayer | 12,898,417 | 7.9 |
| Pfizer | 12,739,444 | 7.8 |
| AstraZeneca | 11,293,007 | 6.9 |
| Amgen | 8,897,356 | 5.4 |
| Boehringer Ingelheim | 8,209,882 | 5 |
| Bristol Myers Squibb | 7,570,771 | 4.6 |
| AbbVie | 7,232,153 | 4.4 |
| MSD | 7,123,710 | 4.3 |
| Sanofi | 7,078,410 | 4.3 |

**Company–specialty patterns**

Payments were not evenly distributed across companies. In five specialties, where one or two products have been marketed, one company was responsible for the majority of payments to clinicians. This was the case for palliative medicine (Menarini, A$ 226,627, 88% of all payments), pain medicine (Seqirus, A$ 413,918, 86% of payments) and ophthalmology (Bayer, A$4,856,842, 69% of payments) (Table S2).

**Table S2: Specialties with one company making the majority of payments, top 5**

| **Specialty** | **Top company** | **Amount (A$)** | **Share of specialty total (%)** |
| --- | --- | --- | --- |
| Palliative medicine | Menarini | 226,627 | 88 |
| Pain medicine | Seqirus | 413,918 | 86 |
| Ophthalmology | Bayer | 4,856,842 | 69 |
| Sexual health medicine | Gilead Sciences | 733,948 | 69 |
| Paediatric respiratory and sleep medicine | Vertex | 149,527 | 65 |

The dominance of individual companies in certain specialties is consistent with product portfolios in niche areas. For instance, Menarini’s analgesic products likely explain its 88% share in palliative medicine. Seqirus’s role in distributing the opioid analgesic tapentadol (Palexia) may account for its 86% share in pain medicine. Bayer’s anti-VEGF agent aflibercept (Eylea) may underpin its 69% share in ophthalmology. Gilead’s HIV products may explain its dominance in sexual health (69%), while Vertex’s cystic fibrosis CFTR modulators could explain its 65% share in paediatric respiratory and sleep medicine.

**Specialty reach**

Figure 3 in the main manuscript was restricted to specialties with at least 150 fellows. Table S3 presents the unrestricted analysis. This table should be interpreted with caution as proportions may be spuriously elevated due to the presence of minor misclassification errors (accentuated by small denominators).

**Table S3: Specialty reach, ranked by proportion of specialists in each field who received at least one payment between Oct 2015 to Oct 2024 (unrestricted), top 10**

| **Specialty** | **Clinicians paid** | **Total clinicians** | **Proportion (95% CI)** | **Total Payments ($A)** |
| --- | --- | --- | --- | --- |
| Haematology and oncology | 1526 | 1776 | 85.9% (84.2-87.5) | 28,395,112 |
| Rheumatology | 398 | 490 | 81.2% (77.5-84.4) | 11,085,314 |
| Paediatric endocrinology | 48 | 68 | 70.6% (58.9-80.1) | 514,612 |
| Respiratory and sleep medicine | 667 | 962 | 69.3% (66.3-72.2) | 9,912,554 |
| Cardiology | 1191 | 1725 | 69.0% (66.8-71.2) | 16,659,021 |
| Endocrinology | 629 | 949 | 66.3% (63.2-69.2) | 11,217,455 |
| Neurology | 547 | 875 | 62.5% (59.3-65.7) | 9,688,201 |
| Nephrology | 427 | 740 | 57.7% (54.1-61.2) | 4,343,825 |
| Sexual health medicine | 76 | 134 | 56.7% (48.3-64.8) | 1,058,001 |
| Paediatric neurology | 39 | 69 | 56.5% (44.8-67.6) | 585,452 |

** Combined denominator = clinical haematology (RACP) + medical oncology*

** Denominators are from the number of registered practitioners in that specialty in 2023/24*

**Figure S1: Median and spread of individual payment over time**

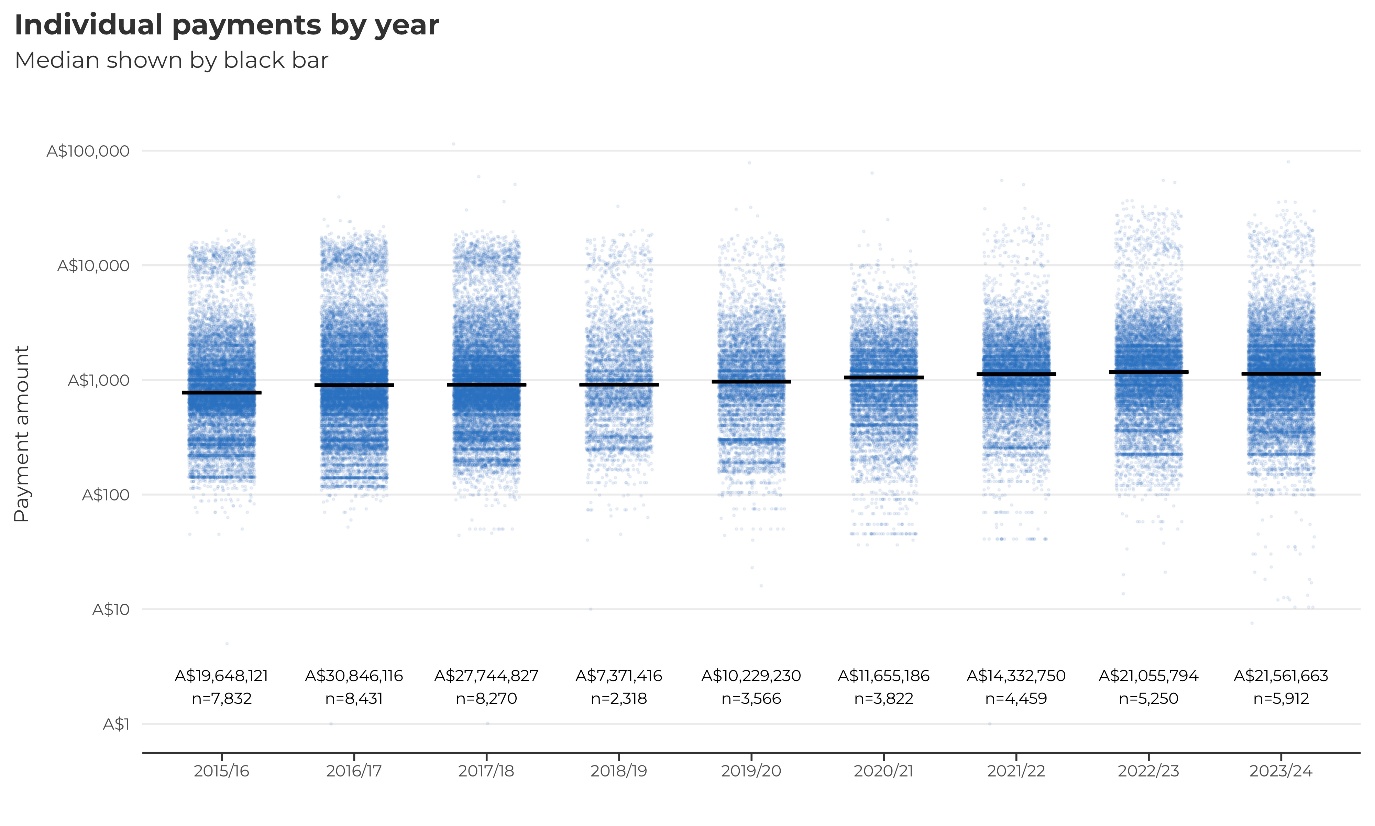

**Figure S2: Lorenz curve demonstrating unequal payment share**

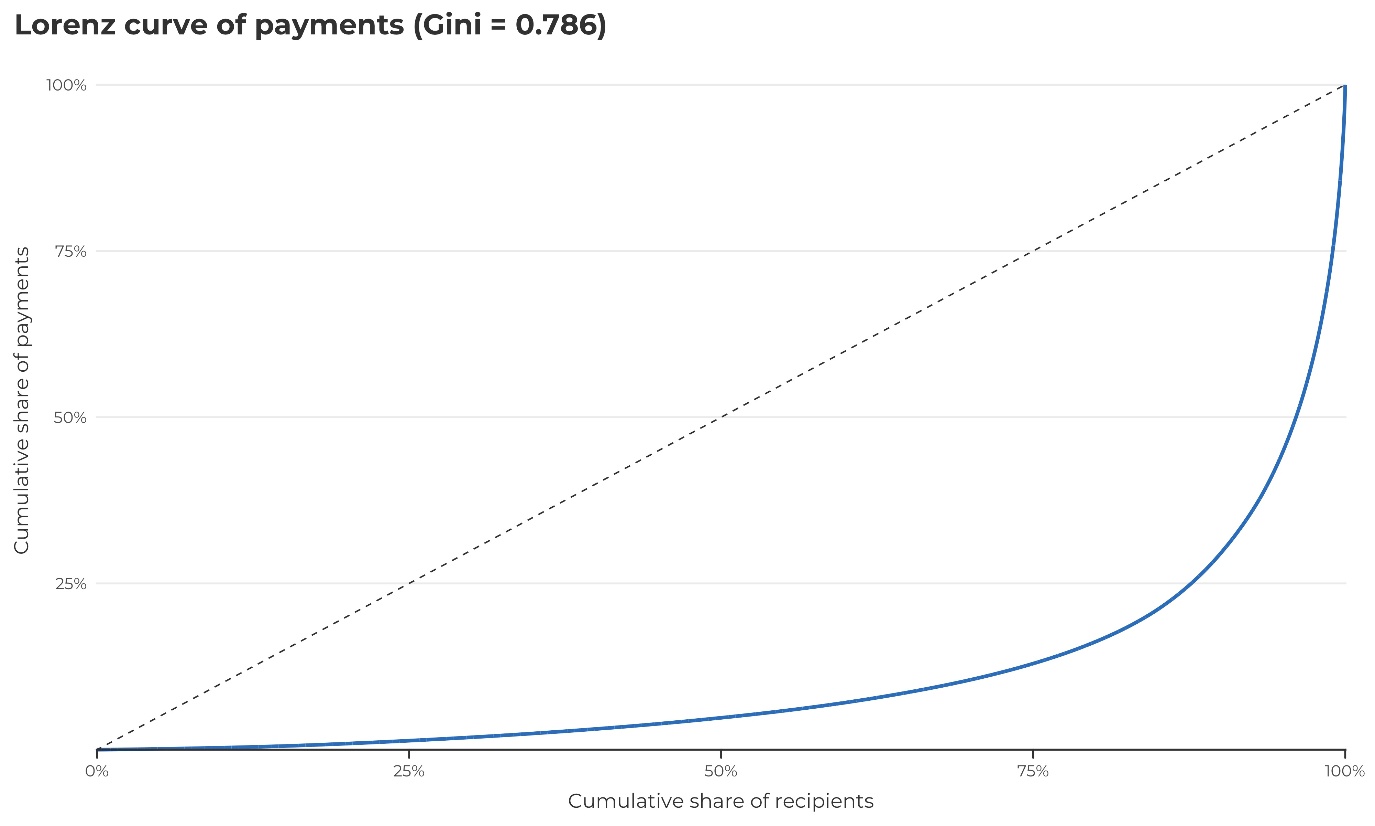

This curve plots the cumulative share of total payments against the cumulative share of recipients, ordered from the smallest to largest total received. The dashed 45° line represents perfect equality. The strong bowing of the observed curve well below this line indicates marked concentration of payments among a small minority of recipients. The associated Gini coefficient of 0.786 (0 = perfect equality; 1 = complete inequality) quantifies this imbalance, showing that most recipients receive comparatively small amounts while a few receive the bulk of total dollars.
